# Correlates of COVID-19 vaccination status among college students

**DOI:** 10.1101/2021.09.15.21263654

**Authors:** Michele Nicolo, Eric Kawaguchi, Angie Ghanem-Uzqueda, Andre E. Kim, Daniel Soto, Sohini Deva, Kush Shanker, Christopher Rogers, Ryan Lee, Frank Gilliland, Jeffery Klausner, Andrea Kovacs, David Conti, Howard Hu, Jennifer B. Unger

**Affiliations:** Department of Population and Public Health Sciences, Keck School of Medicine, University of Southern California, Los Angeles California; Family Medicine, Keck Medicine of USC, Los Angeles, California; Keck School Medicine of USC, University of Southern California, Los Angeles California

## Abstract

**Objectives:** Despite the widespread availability of COVID-19 vaccines in the United States, vaccine hesitancy remains high among certain groups. This study examined the correlates of being unvaccinated among a sample of university students (N=2900) during the spring and summer of 2021, when the campus had been closed for over a year and students were preparing to return to in-person learning.

**Methods:** Students responded to an email invitation and completed electronic surveys. Results. In multivariable logistic regression analyses, students were more likely to be unvaccinated if they were African American, identified with any political affiliation other than Democrat, were undergraduates or international students, had not traveled outside the Los Angeles during the pandemic, and/or had previously been ill with COVID-19.

**Conclusion:** Findings indicate that culturally resonant educational interventions, and possibly vaccine requirements, are needed to promote vaccination among university students.

## Introduction

The recent slowing of COVID-19 vaccination among adults suggests vaccine hesitancy in some individuals and groups.[1] Reasons for vaccine hesitancy include disparities in access to vaccines and accurate information, political influences, concern over adverse health effects including infertility, and mistrust of the medical community [2–5].

Given the rising trend in COVID-19 cases, the increased risk posed by the reopening of college campuses, low vaccine uptake among young adults, the dynamic nature of this pandemic, and the fact that young adult vaccination is a critical component needed to reduce morbidity and mortality, research on vaccine hesitancy among college students is needed. Successful health communication campaigns to encourage vaccination among this group will depend on a thorough understanding of the epidemiology of vaccine hesitancy. To investigate college student vaccine hesitancy, this study examined characteristics associated with being unvaccinated among university students during the spring and summer of 2021.

## Methods

### Participants

Participants were students at the University of Southern California (USC) in Los Angeles, California. Students were eligible if they were currently enrolled at USC, were at least 18 years of age, and provided informed consent.

### Procedure

The USC Institutional Review Board approved the study. The study was advertised on university websites. Emails were sent to all undergraduate and graduate students inviting them to participate in a brief COVID-19 survey. Interested students clicked on a link in the email, provided informed consent electronically, and completed the online survey. Survey responses were collected from 4/29/2021 to 07/12/2021, with 86.3% of the surveys completed in April and May 2021. After removing duplicate and incomplete surveys, the sample size was 2900.

### Measures

Demographic variables included self-identified race and ethnicity (White; Asian/Asian American; Black/African American; Hispanic/Latinx; multicultural; or other), sex, degree status (undergraduate versus graduate student), international student status (yes/no), and political affiliation (Democrat, Independent, Republican, something else). COVID-19 history was self-reported and categorized as “yes” or “no”. Additional questions asked about housing, degree program, recent travel outside of Los Angeles, COVID-19 knowledge, and attitudes and compliance with prevention behaviors including masking. The outcome variable was self-reported vaccination status at the time of the survey.

### Data analysis

Univariate logistic regression models were run to select variables for inclusion in the final models. Variables that were significantly associated with vaccination status at p<0.05 were included in a final multivariable model. Adjusted odds ratios and 95% confidence interval associated with vaccination status are reported.

### Results

Among the 28,412 students who were sent invitation emails, 2,900 students completed the survey and were included in the analysis. Table 1 shows the demographic characteristics of the sample. The demographic characteristics of the sample were similar to those of the USC student population, although the sample was overrepresented by Asians and women and underrepresented by Latinx/Hispanic and international students. Most participants (82.9%) self-reported that they were vaccinated at the time of the survey.

**Table 1.** Characteristics associated with the likelihood of not receiving the COVID-19 vaccine among university students using an adjusted binary logistic regression model. (N=2900)

| Characteristic | N % | Odds Ratio | 95 % Confidence Interval |
| --- | --- | --- | --- |
| <u>Race/ethnicity</u> |  |  |  |
| Asian or Asian American | 997 (34.4) | Reference | Reference |
| Black or African American | 142 (4.9) | 1.14 | 0.86, 1.51 |
| Hispanic/Mexican/Spanish/Latinx | 218 (7.5) | 1.88 | 1.20, 2.96 |
| Multicultural or other | 682 (23.5) | 1.49 | 0.99, 2.28 |
| White | 861 (29.7) | 0.97 | 0.72, 1.30 |
| <u>Sex</u> |  |  |  |
| Female | 1853 (63.9) | Reference | Reference |
| Male |  | 0.95 | 0.77, 1.18 |
| <u>Student Status</u> |  |  |  |
| Undergraduate | 1384 (47.8) | Reference | Reference |
| Graduate | 1514 (52.2) | 1.39 | 1.13, 1.72 |
| <u>International student</u> |  |  |  |
| Yes | 188 (6.7) | Reference | Reference |
| No | 2629 (93.3) | 2.80 | 1.97, 3.98 |
| <u>Traveled in/out of Los Angeles</u> |  |  |  |
| Yes | 1882 (65.8) | Reference | Reference |
| No | 978 (34.2) | 0.77 | 0.62, 0.96 |
| <u>Political Affiliation</u> |  |  |  |
| Democrat | 1876 (64.7) | Reference | Reference |
| Independent | 563 (19.4) | 2.31 | 1.81, 2.95 |
| Republican | 178 (6.1) | 1.56 | 1.03, 2.35 |
| Something else | 283 (9.8) | 2.31 | 1.63, 3.27 |
| <u>Had COVID-19</u> |  |  |  |
| Yes | 622 (21.4) | Reference | Reference |
| No | 2278 (78.6) | 1.98 | 1.56, 2.50 |

Table 1 shows the odds ratios and 95% confidence intervals from the multivariable analysis of characteristics associated with the likelihood of being unvaccinated. African Americans had higher odds of being unvaccinated compared to Non-Hispanic White participants (OR=1.88, 95% CI: 1.20, 2.96). Undergraduate students (OR= 1.39, 95% CI: 1.13. 1.72) and international students (OR=2.80, 95% CI: 1.97, 3.98) had higher odds of being unvaccinated when compared to graduate students and domestic students, respectively. Participants reporting previous COVID-19 illness (OR= 1.98, 95% CI: 1.56, 2.50) had higher odds of being unvaccinated compared to students reporting no history of having COVID-19. Identifying political affiliation as Independent (OR=2.31, 95% CI:1.81, 2.95), Republican (OR=1.56, 95% CI: 1.03, 2.35) or something else (OR= 2.31, 95% CI: 1.63, 3.27) were associated with greater odds of being unvaccinated compared to Democrats. Students who had traveled out of the Los Angeles area during the pandemic were more likely to be vaccinated (OR=0.77, 95% CI: 0.62, 0.96) compared to students who remained in Los Angeles. No other variables were associated with being unvaccinated.

## Discussion

This study identified correlates of being unvaccinated for COVID-19 among a large sample of university students during the spring and summer of 2021, when the university had been operating completely online for over a year and before implementation of a vaccination policy for campus reopening. At the time of survey completion, a large percentage of student participants (82.9%) reported that they had already received the COVID-19 vaccine. This increased to 94.2% after a vaccine mandate was announced later in the summer.

The significant risk factors for being unvaccinated included African American race, all political affiliations except Democrat, previous history of COVID-19 infection, staying in the Los Angeles area during the pandemic, and being an undergraduate or international student. Consistent with previous studies [23-32], African Americans were especially likely to be unvaccinated. Low vaccination uptake among African Americans has been attributed a history of racial discrimination and injustice resulting in distrust of the medical community, and disparities in vaccine access. Culturally relevant health communications from trusted spokespeople in the African American community could potentially increase vaccination among this group.[6]

Students who identified as Democrats were more likely to be vaccinated than those with other political affiliations. The COVID-19 vaccine was developed and received emergency authorization in the U.S. during a time of political tension.[5, 7–9] Other studies of college students and other adults have found that conservative political ideology is associated with vaccine hesitancy and distrust.[10] Interventions are needed to counter distrust of science and government.

Previous history of COVID-19 infection was associated with being unvaccinated. Students who had had COVID-19 might have believed that they already had immunity and did not need the vaccine. However, recent studies have shown that vaccination prevents reinfection and severe disease among people who have recovered from COVID-19. [11]

Previous studies have documented vaccine hesitancy and skepticism among university students, including low perceived risk of serious illness, mistrust of the vaccine development process, perception that the vaccine was developed too quickly, doubts about the safety of the vaccine, and lack of information [32,33]. However, previous studies were limited by small samples of students enrolled in specific degree programs. This study extends the findings of previous studies by employing a large, diverse sample consisting of undergraduate and graduate students across academic disciplines, during a pivotal time period when they were making decisions about whether to get vaccinated to prepare for the return to in-person learning in the fall of 2021.

### Limitations

This analysis was based on a non-random sample of university students who responded to an online survey. Findings may not generalize to the larger student population or to other universities. Future studies are needed to identify correlates of vaccination and vaccine hesitancy among students who chose not to participate. Vaccination status was self-reported.

## Conclusions

This study identifies subgroups of university students who were vaccine-hesitant prior to the start of the 2021-2022 academic year. Although some of these students subsequently got vaccinated after a vaccine policy was implemented, this study identifies groups that potentially need more education and outreach to promote vaccination. Understanding factors contributing to vaccine hesitancy provide the necessary knowledge of how to target education campaigns across college and university campuses, presenting an opportunity to collaborate with students and leadership to build reliable, evidence-based resources to face future pandemics.

## Data Availability

Data available upon request

